# Medial temporal lobe Tau-Neurodegeneration *mismatch* from MRI and plasma biomarkers identifies vulnerable and resilient phenotypes with AD

**DOI:** 10.1101/2025.08.17.25333859

**Authors:** Xueying Lyu, Nidhi S Mundada, Christopher A. Brown, Niyousha Sadeghpour, Emily McGrew, Long Xie, Yue Li, Anika Wuestefeld, Michael Tran Duong, Philip Cook, James Gee, Ilya M Nasrallah, Alice S. Chen-Plotkin, Leslie M. Shaw, Dawn Mechanic-Hamilton, L.E.M. Wisse, Paul A Yushkevich, Sandhitsu R Das, David A Wolk, the Alzheimer’s Disease Neuroimaging Initiative

## Abstract

While tau pathology is closely associated with neurodegeneration in Alzheimer’s disease (AD), our prior work using multi-modality imaging revealed that mismatch between tau (T) and neurodegeneration (N) may reflect contributions from non-AD processes. The medial temporal lobe (MTL), an early site of AD pathology, is also a common target of co-pathologies such as limbic-predominant age-related TDP-43 encephalopathy neuropathologic change (LATE-NC), often following an anterior–posterior atrophy gradient. Given the susceptibility of MTL to co-pathologies, here we explored T-N mismatch specifically within MTL using plasma ptau_217_ and MTL morphometry for identifying vulnerabilities and resilience in cognitively impaired or unimpaired AD patients.

We parcellated the MTL into 100 spatially contiguous segments and calculated their T-N mismatch using plasma ptau_217_ as a measure for T and thickness as a marker of N. Based on these mismatch profiles, we clustered 447 amyloid-positive individuals from ADNI cohort into data-driven T-N phenotypes. We characterized the T-N phenotypes by examining their cross-sectional and longitudinal atrophy both within the MTL and across the whole brain, as well as cognitive trajectories. This framework was replicated in an independent cohort and finally translated to a real-world clinical sample of 50 patients undergoing anti-amyloid therapy.

Clustering identified three T-N phenotypes with different MTL T-N mismatch profiles, atrophy patterns, and cognitive outcomes, despite comparable AD severity. The “canonical” group, characterized by low T-N residuals (N ∼ T), showed AD-like neurodegeneration patterns. The “vulnerable” group, characterized by disproportionately greater neurodegeneration than tau (N > T), showed atrophy primarily in the anterior MTL that extended into temporal-limbic regions, both in cross-sectional and longitudinal analyses. This group also exhibited neurodegeneration that preceded estimated tau onset and experienced faster cognitive decline across multiple domains, aligning with the typical characteristics of mixed LATE-NC with AD. In contrast, the “resilient” group (N < T) showed minimal atrophy and preserved cognitive function. These phenotypes were reproducible in an independent research cohort. Importantly, in a feasibility study applying the model developed from ADNI to a clinical cohort of patients receiving lecanemab, we identified vulnerable individuals with LATE-like atrophy patterns. This highlights its potential utility for identifying individuals with co-pathology in clinical settings.

Our findings demonstrate that T-N mismatch within MTL using MRI and plasma biomarkers can reveal AD groups with varying vulnerability/resilience, with the vulnerable group displaying structural and cognitive outcomes suggestive of LATE-NC. This approach offers a cost-effective strategy for clinical trial stratification and precision medicine for AD therapeutics.

## Introduction

The recent Alzheimer’s Association Workgroup revised diagnostic criteria build upon the foundation of the previously described ATN framework, which was a first attempt at a biological definition of AD.^1,2^ Based on these criteria, amyloid-β plaques (A) and neurofibrillary tangles (NFT) are the defining pathologies of Alzheimer’s disease (AD)^1,2^ that mark its presence and stage, respectively. However, AD is a heterogenous condition commonly associated with additional pathologies, such as TDP-43, alpha-synuclein, and cerebrovascular disease. Detecting the presence of these co-pathologies and characterizing their role in disease progression is a critical knowledge gap in the context of emergent disease-modifying therapies^3,4^. The lack of specific and quantitative molecular biomarkers of these co-pathologies (e.g. TDP-43) ^5–10^ is a major barrier to such characterization.

The medial temporal lobe (MTL)^11^ is a site of early neurodegeneration in AD. Most notably, tau (T) accumulation in the cerebral cortex typically begins in Brodmann area 35 (BA35) and the entorhinal cortex and then spreads to the hippocampus proper^12–16^. The accumulation of T drives neurodegeneration (N) within the MTL, making it a clinically important region to aid in early AD diagnosis. Nevertheless, the N in MTL is not solely driven by T, as the region also serves as a critical locus of early pathology for co-pathologies in addition to AD. Limbic-predominant age-related TDP-43 encephalopathy neuropathologic change (LATE-NC) ^10,17^, which is present in up to 50% of patients with AD^18^, also involves MTL structures, with involvement of the amygdala and then hippocampus and entorhinal cortex defining Stage 1 and 2 of this condition, respectively^19,20^. Moreover, previous work has found that there is an anterior-to-posterior gradient of involvement of LATE-NC along the long axis of the MTL, most prominently affecting anterior MTL regions and extending into anterior medial temporal polar regions^19,21^. Additionally, other pathologies, such as alpha-synuclein and vascular disease, also contribute to MTL structural changes, although, unlike ADNC and LATE-NC, MTL is not thought to be the epicenter for these pathologies ^19,22–24^. Therefore, characterizing the heterogeneity within the MTL is crucial for advancing precision medicine approach in AD therapeutics.

Despite the close link between the burden of T and downstream N^12,25–28^ in AD, there is still considerable variability in this relationship across individuals. One potential source of this variability is the presence of co-pathologies which may have independent effects on measures of neurodegeneration. In our prior work^29–31^, we quantified this variability by determining the degree of *mismatch* in the relationship between T-N across the whole brain using either MRI or ^18^F-fluorodeoxyglucose PET and tau PET as a surrogate of N and T, respectively. By examining maps of residuals between markers of T and N across the brain and using clustering techniques to find groups with characteristic T/N residual patterns, we identified distinct T-N *mismatch* phenotypes. Based on both in vivo and ex-vivo studies^29–31^, we found that these T-N phenotypes were enriched with specific co-pathologies (e.g. TDP-43 and vascular disease).

Given the susceptibility of MTL to multiple pathologies, particularly LATE-NC^10,19,24^, in this work, we specifically investigated the T-N *mismatch* within this region by integrating structural MRI as a marker of N and a plasma biomarker as a marker of T. Plasma ptau_217_, a non-invasive yet highly specific AD biomarker, has been shown to have high sensitivity for the presence of cerebral amyloid-β as measured by amyloid-β PET, but to also be related to tau burden as measured by tau PET^32–34^. While tau PET may be more tightly linked to NFT burden and provides spatial information, plasma ptau_217_ is a more practical and cost-effective measure for clinical practice. As MRI is also routinely acquired in clinical practice, especially in the course of anti-amyloid treatment, in the current analysis we set out to use plasma ptau_217_ as a measure of T and MTL thickness, combined with amygdala volume, as a surrogate of N. To study T-N mismatch at a finer spatial resolution, we divided the MTL into 100 segments of approximately equal size, referred to as “super-points.” This more granular representation was designed to better capture spatial heterogeneity in T-N mismatch within the MTL than using larger anatomically defined subregions (e.g. CA1, ERC etc).

Using this simplified T-N framework, **(1)** we used a data-driven clustering approach to cluster amyloid-β-positive (A+) individuals with and without cognitive impairment from ADNI into a discrete set of groups based on similarity of T-N *mismatch* patterns across the 100 MTL super-points and amygdala. Based on our prior whole brain T-N mismatch studies^29–31^, here we anticipated identifying distinct T-N groups with negative residuals (N > T; vulnerable) or positive residuals (N < T; resilient) in MTL**. (2)** We examined cross-sectional and longitudinal atrophy trajectories in the MTL and across the whole brain, along with cognitive profiles, in these T-N groups. We hypothesized that the vulnerable group would be enriched in LATE and would exhibit LATE-like biomarker patterns, including an anterior-to-posterior gradient of atrophy in the MTL and a more impaired cognitive profile, predominantly in memory. **(3)** We further examined relationships of cognitive and neurodegenerative trajectories anchored to a time of tau positivity onset estimated using the Statistical Inference for Longitudinal Analysis (SILA) approach^35^ . **(4)** Next, we replicated this approach in an independent cohort to assess its reproducibility. **(5)** Finally, we applied the method to a real-world clinical cohort of patients treated with anti-amyloid-β therapy to evaluate its translational potential.

## Methods

### Participants

Data used in the preparation of this article were obtained from the Alzheimer’s Disease Neuroimaging Initiative (ADNI) database (adni.loni.usc.edu). The ADNI was launched in 2003 as a public-private partnership, led by Principal Investigator Michael W. Weiner, MD. The primary goal of ADNI has been to test whether serial magnetic resonance imaging (MRI), positron emission tomography (PET), other biological markers, and clinical and neuropsychological assessment can be combined to measure the progression of mild cognitive impairment (MCI) and early Alzheimer’s disease (AD).

Participants were selected from ADNI based on the following criteria: (1) amyloid-positive (A+) status that were either cognitively unimpaired, mild cognitive impairment (MCI) or dementia, (2) availability of a pair of plasma ptau_217_ and T1-weighted MRI scans acquired within one year of each other, (3) successful completion of T1-weighted MRI image processing, and (4) passing quality control (QC) for image processing. We identified 469 A+ patients from ADNI with a pair of plasma ptau_217_ and T1-weighted MRI scan within one year apart, based on data accessed as of March 2025. After excluding 11 individuals due to image processing failures (see below section MTL thickness and subregional volume estimation) and another 11 for failed quality control for segmentation, a total of 447 A+ individuals were included in the analysis. The average time difference between the MRI scan and ptau_217_ measurement was 0.84 ( ± 1.97) months. Detailed clinical characteristics of this cohort including age, sex, CDRSB, ptau_217_, are provided in Table S1. We additionally included 123 amyloid-β-negative (A−) cognitively unimpaired individuals from ADNI as a control group. The average age of this group was 74.6 years (± 6.7), comprising 51 females and 72 males.

To form an independent replication cohort, we included 108 A+ individuals from the University of Pennsylvania Alzheimer’s Disease Research Center (Penn ADRC), applying the same patient selection criteria as was used for ADNI. Data were collected from April 2016 to January 2025. The average interval between MRI and plasma ptau_217_ measurement was 3.6 ± 3.4 months. Clinical details for the Penn ADRC cohort are provided in Table S1.

Finally, as a clinical translation cohort, we included 50 patients with MCI or mild dementia due to AD from the University of Pennsylvania Anti-Amyloid Therapy Monitoring (ATM) program, all of whom have received lecanemab. All patients underwent MRI and plasma biomarker measurement prior to starting treatment, with data collected between May 2024 and January 2025. All participants provided informed consent in accordance with protocols approved by the University of Pennsylvania Institutional Review Board. The average interval between ptau_217_ and MRI in this group was 4.3 (±2.4) months. Clinical details for ATM cohorts are also summarized in Table S1.

### Image acquisition and processing

#### Image acquisition

The image acquisition methods for ADNI have been previously described^29,30^. Briefly, The T1-weighted MRI scans were acquired with a spatial resolution of 1.0 × 1.0 × 1.0 mm³ or 1.0×1.0×1.2mm^3^.

Structural MRI data for the Penn ADRC study were collected on a Siemens Prisma 3 tesla (T) scanner using a 3D T1-weighted MPRAGE sequence, featuring the following parameters: TR = 2400 ms, TE = 2.24 ms, TI = 1060 ms, flip angle = 8°, field of view = 256 × 240 × 167 mm³, and voxel dimensions of 0.8 × 0.8 × 0.8 mm³.

For the ATM program, T1-weighted structural MRI scans were acquired using Siemens 1.5T (MAGNETOM Sola, Avanto) and 3T scanners (Prisma, Skyra, Vida). Most scans employed a 3D MPRAGE or similar sequence (TR ≈ 2300 ms, TE ≈ 2.9 ms, TI ≈ 900 ms, flip angle ≈ 8–9°, voxel size ≈ 0.8×0.8×0.8 mm³ or 1.0×1.0×1.0 mm³).

#### Amyloid-β status determination

Amyloid-β status for ADNI participants was determined based on amyloid-β PET imaging, using tracer- and reference-specific SUVR thresholds by UC Berkeley. Given most of participants from ADNI had amyloid PET, we used amyloid-β PET imaging for amyloid status determination which still remains the gold-standard for amyloid classification. For the Penn ADRC cohort, amyloid-β status was based on visual reads of amyloid PET scans from a neuroradiologist with nuclear medicine training when available. For Penn ADRC participants without amyloid-β PET, amyloid-β status was determined using the ptau217/Aβ42 ratio, with a cutoff of >0.0055 indicating amyloid-β positivity. This threshold corresponds to 95% sensitivity and approximately 90% overall classification accuracy against amyloid PET^36^.

#### MTL thickness and subregional volume estimation

MTL thickness was derived through a previously-validated automatic pipeline^37^. First, the MTL was segmented into subregions (anterior and posterior hippocampus, amygdala, entorhinal cortex, parahippocampal cortex, BA35 and BA36) from T1-weighted MRI using the *Automated Segmentation of Hippocampal Subfields* algorithm/software (ASHS)^38–40^, and volumes obtained. Intracranial volume (ICV) was also estimated by ASHS. Next, thickness estimation was performed using the *Cortical Reconstruction for ASHS* pipeline (CRASHS)^37^. Detailed description of CRASHS has been outlined here^37^. Briefly, it generates participant-specific gray/white, pial, and mid-surfaces as triangular meshes for the MTL as the input for estimating thickness. To allow group-level morphometric analysis, individual MTL surfaces were aligned to a population template using diffeomorphic surface registration.

#### MTL super-points parcellation

We developed a super-point partitioning approach to further parcellate the MTL into 100 bilateral, spatially contiguous segments of approximately equal size, referred to as “super-points,” for cortical thickness analysis. This was done to allow more granular T-N mismatch analyses than using larger anatomically defined subregions. We defined the super-points once on CRASHS surface template, which is derived from inflated MTL mid-surfaces of a set of individuals on the AD continuum and represents a smooth “average” MTL shape^37^. Our prior work^29–31^ demonstrated stable clustering performance with 100 input features, we therefore selected this number to balance spatial resolution with stability. The number of super-points assigned to each subregion was approximately proportional to its surface area so that larger regions were represented by more partitions. Within each region, we constructed a triangle adjacency graph and applied PyMetis-based clustering^41^ to generate spatially contiguous super-points, ensuring partitions remained confined within the anatomical boundaries of each region. Using the vertex-wise MTL thickness maps generated by CRASHS, we calculated the mean thickness for each super-point for each participant. This process produced MTL thickness estimates for 100 super-points.

#### Whole brain cortical thickness processing

In addition to MTL thickness, T1-weighted MRI scans were processed using the ANTs cortical thickness pipeline^42^, which includes intensity inhomogeneity correction and tissue segmentation. A multi-atlas segmentation approach^43^ was used to parcellate the scans into cerebellar, cortical, and subcortical regions of interest (ROIs). Volumetric thickness maps for each participant were then generated using the DiReCT cortical thickness estimation method^44^.

Longitudinal cortical thickness measurement used the ANTs longitudinal pipeline^45^. This method creates an unbiased within-participant template from all timepoints to allow consistent alignment across scans and reduce bias. Cortical thickness is then estimated using tissue segmentation in the native space.

### Plasma ptau_217_

All included individuals for T-N *mismatch* analyses had plasma ptau_217_ samples. All samples were analyzed on the Fujirebio Lumipulse G1200 analyzer at either the University of Pennsylvania ADNI Biomarker Core (ADNI, Penn ADRC, ATM) or University of Indiana Biomarker Assay Lab (ADNI). Within the ADNI cohort, 291 samples were analyzed at the University of Pennsylvania and 156 at the University of Indiana. Tau positivity was defined as plasma ptau_217_ > 0.3175. This threshold was determined in a previously described cohort^46^ with both Tau-PET and plasma measures available^46^ to identify the ptau_217_ value with the maximal Youden index for identifying individuals exceeding a Tau-PET threshold > 97.5th percentile of cognitively unimpaired, amyloid negative (A-) individuals.

### Cognitive testing

Global clinical impairment was assessed using the Clinical Dementia Rating Sum of Boxes (CDRSB) for both ADNI and Penn ADRC. Domain-level cognitive performance was derived from the ADSP Phenotype Harmonization Consortium’s composite scores dataset (ADSP_PHC_COGN_domain_wise_score) available through ADNI. This harmonized dataset provides standardized z-scores for cognitive domains, including memory, executive function, and language.

### T-N *mismatch* modeling

To capture spatial patterns of T-N *mismatch* within the MTL, we used cortical thickness from the 100 super-points as a measure of N and plasma ptau_217_ as a measure of T. For each super-point, we performed robust linear regression^47^ between its corresponding thickness and ptau_217_ with Tukey’s bisquare function to downweight the influence of outliers. We then extracted the residuals as a measure of T-N mismatch. We also performed a separate regression of amygdala volume on ptau_217_, controlling for ICV to obtain an additional residual reflecting mismatch in the amygdala. Residuals from both the super-points and amygdala were discretized into three categories (+1, 0, −1) based on whether they were more than 0.5 standard deviations above or below the regression line. Further, the discretized residual for amygdala in each hemisphere was weighted seven times that of a single super-point residual, as the amygdala volume is approximately 7 times the volume of a single super-point. The combined residual profiles were then clustered using Ward’s hierarchical clustering^48^ based on Euclidean distance to derive data-driven T-N mismatch phenotypes. The number of clusters was guided by visual inspection of the dendrogram (Figure S1) and informed by our prior work^29–31^, which identified three MTL-involved groups at the whole-brain level, suggesting that a three-cluster solution may be optimal. This modeling approach was used to generate T–N groups in the ADNI cohort for the main analyses and replicated in an independent cohort from the Penn ADRC.

### Assigning Clinical Patients to ADNI-Derived T–N Phenotypes

As a proof of concept for clinical practice, we applied the T–N mismatch model trained on the ADNI cohort to an independent clinical dataset of patients undergoing anti-amyloid-β therapies (ATM cohort). For each patient in ATM, T–N residuals were computed at super-points and weighted amygdala regions using ADNI-derived regression models. These residuals were then binarized using a threshold of ±0.5 standard deviations from the corresponding ADNI distribution. Each ATM patient was then matched to one of the three ADNI-derived T–N phenotypes by identifying the cluster centroid with the closest Euclidean distance in the binarized residual space. To facilitate broader application, we released a user-friendly software package for implementing the mismatch approach and individual T-N phenotypic classification (https://github.com/xueying-lyu/mismatch).

### Imaging signature of LATE-NC

A prior postmortem study^19^ of MTL comorbidities demonstrated that the thickness ratio between the entorhinal cortex and parahippocampal cortex (ERC/PHC) was reduced in AD patients with TDP-43 co-pathology. Based on this, we used the ERC/PHC ratio as an imaging surrogate for LATE-NC and compared this metric across the T-N groups in this work.

### Sampled Iterative Local Approximation (SILA)

To anchor longitudinal analyses within the disease time course, we used SILA^49^ to estimate individuals’ Tau onset age. Briefly, SILA uses short-term longitudinal follow-up data to approximate a single trajectory for change in a given biomarker over disease duration. This then allows for estimation of the age at which a biomarker becomes abnormal from any single timepoint. For our study, we combined data from 397 A+ individuals in the ADNI and Penn ADRC cohorts with longitudinal ptau_217_ available to generate a p-tau_217_ trajectory with a threshold of 0.3175 for tau positivity (as described above). We then used this trajectory to estimate tau onset age for each participant in our dataset.

### Statistical modeling and analyses

#### Analyses overview

We first performed group comparisons between mismatch groups in the primary ADNI cohort to evaluate cross-sectional and longitudinal structural differences as well as differences in clinical and cognitive trajectories. We then replicated cross-sectional thickness comparisons and longitudinal cognitive impairment analyses in the Penn ADRC cohort. Finally, we performed vertex-wise cortical thickness comparisons in the MTL for the clinical ATM cohort. Details of these analyses are provided below.

#### General statistical analyses

All general statistical analyses were performed using R (v4.5). Between-group comparisons of continuous variables, including age, ptau_217_, imaging features, and cognitive scores, were conducted using general linear models, with group as the independent variable and relevant covariates included for adjustment. The comparisons were two-sided. Ordinal variables, such as clinical diagnosis, were compared using the Kruskal–Wallis test^50^.

#### Cross-sectional thickness comparisons

##### MTL surface vertex-wise thickness comparisons

We performed vertex-wise comparisons of MTL thickness across T-N groups using a general linear model as implemented in meshglm program (https://github.com/pyushkevich/cmrep). Prior to modeling, participant-level MTL thickness maps were smoothed using surface-based diffusion smoothing with a Gaussian kernel (σ ≈ 3 mm) applied on the mesh. For visualization, we fitted the CRASHS template to an ASHS segmentation of the MNI template^51^ and projected thickness map into this geometry, which more closely reflects typical MTL anatomy than the inflated CRASHS template. At each vertex of the template surface mesh, thickness was modeled as the dependent variable, group membership as the main factor of interest, and age, ptau_217_, and sex as covariates. We controlled for p-tau_217_ to remove reduce effects of differences in tau severity between groups. For comparisons between T-N groups and cognitively unimpaired individuals, the model included age and sex as covariates. Group contrasts were computed at each vertex, and t-statistics were derived. To correct for multiple comparisons, we applied permutation testing with 10000 iterations using threshold-free cluster enhancement (TFCE)^52^ with parameters E = 0.5, H = 2, and Δh = 0.1, yielding family-wise error rate^53^ (FWER)-corrected P-values at each vertex.

##### Whole brain voxel-wise thickness comparisons

Additionally, voxel-wise comparisons of whole-brain gray matter thickness across groups were performed with FSL’s randomize tool using TFCE for 10000 permutations, controlling for age, sex and ptau_217_. For comparisons between T-N groups and cognitively unimpaired individuals, age and sex were included as covariates. Similarly, the p-values were corrected by FWER.

#### Longitudinal thickness analyses

##### MTL vertex-wise atrophy

We performed vertex-wise linear mixed-effects modeling^54^ using lme4 package in R with longitudinal MTL thickness as the dependent variable, group*time interaction as the fixed effect, and participant as the random effect, controlling for age, ptau_217_, and sex. We restricted the time from baseline to a ±5 year window. Among 447 ADNI participants, 370 had longitudinal MRI scans, with an average of 4.08 ± 1.80 scans per participant over an average follow-up span of 3.49 ± 1.96 years. The 123 A– CU controls from ADNI had 3.3 ± 1.34 timepoints over 4.07 ± 1.85 years of follow-up. Differences in thickness changes across groups were determined by testing the significance of the time * group interaction term. Comparisons with CU controls were adjusted for age and sex. Group-wise T-statistics and FDR-corrected P-values were computed.

##### Whole brain voxel-wise atrophy

To assess whole-brain longitudinal atrophy differences between groups, we applied voxel-wise linear mixed-effects modelling using lmerNIFTI^55^. We included time points from baseline between -5 to 5 years. Due to the computational limitation, for participants with more than two scans, we required that each scan be at least one year apart. Of the 370 ADNI T-N patients with longitudinal MRI scans, 7 failed whole-brain longitudinal processing, resulting in 363 patients with a mean of 3.03 ± 1.06 timepoints over an average follow-up period of 3.36 ± 2.01 years. The 123 A– cognitively unimpaired controls from ADNI had 2.95 ± 0.96 timepoints over 4.02 ± 1.83 years of follow-up. Longitudinal cortical thickness was modeled as the dependent variable, with group*time interaction as the fixed effect, and participant as the random intercept, controlling for age, ptau_217_, and sex. For the comparisons of groups to CU controls, we only controlled for age and sex. Differences in thickness changes across groups were determined by testing the significance of the time * group interaction term. Resulting p-values were corrected for multiple comparisons using FDR.

##### Longitudinal atrophy preceding T+

For analyses focused on the period preceding estimated Tau positivity (T+; see above section *<u>Estimated Tau onset age using SILA</u>*), we restricted the data to the interval from -15 to 0 years relative to estimated T+ age. Among the included 447 A+ patients from ADNI, there were 172 A+ individuals with longitudinal MRI scans before estimated tau onset, with a mean of 2.84 ± 1.04 time points over 3.05 ± 2.00 years. We again conducted both longitudinal MTL and whole brain voxel-wise analyses based on linear mixed effect models following the same procedures described above. Similarly, the models included the interaction between time-from-T+ and group as the fixed effect, but with age at estimated T+ and sex as covariates.

#### Longitudinal cognitive impairment analyses

##### Modeling longitudinal cognitive trajectories relative to T+

Cognitive trajectories were analyzed using linear mixed-effects models^54^, with cognitive scores as the outcome. We included cognitive timepoints within a ±5-year window relative to the estimated T+ time (see above section *<u>Estimated Tau onset age using SILA)</u>*. The models included fixed effects for time*group interaction (using b-spine with 2 degrees of freedom), as well as covariates for age at estimated T+, sex, and education. A random intercept was included to account for within-participant variability. Differences in cognitive decline across groups were determined by testing the significance of the time * group interaction term. All statistical tests were two-sided. The p-values were Bonferroni-corrected for multiple comparisons. We also repeated the same modeling procedure with time anchored to baseline instead of estimated T+, including ptau_217_, age at baseline, sex, and education as covariates.

## Results

### T-N mismatch groups

In order for our T-N mismatch measurements to be meaningful, a significant association between T and N must be observed across most of the MTL. To assess this, we first examined vertex-wise associations between plasma ptau_217_ and MTL regional thickness among 447 A+ individuals from ADNI (Figure 1A). Level of ptau_217_ was significantly associated with reduced cortical thickness throughout the entire MTL area, with the strongest effects observed in hippocampal and anterior extrahippocampal areas, after adjusting for age and sex.

**Figure 1.**
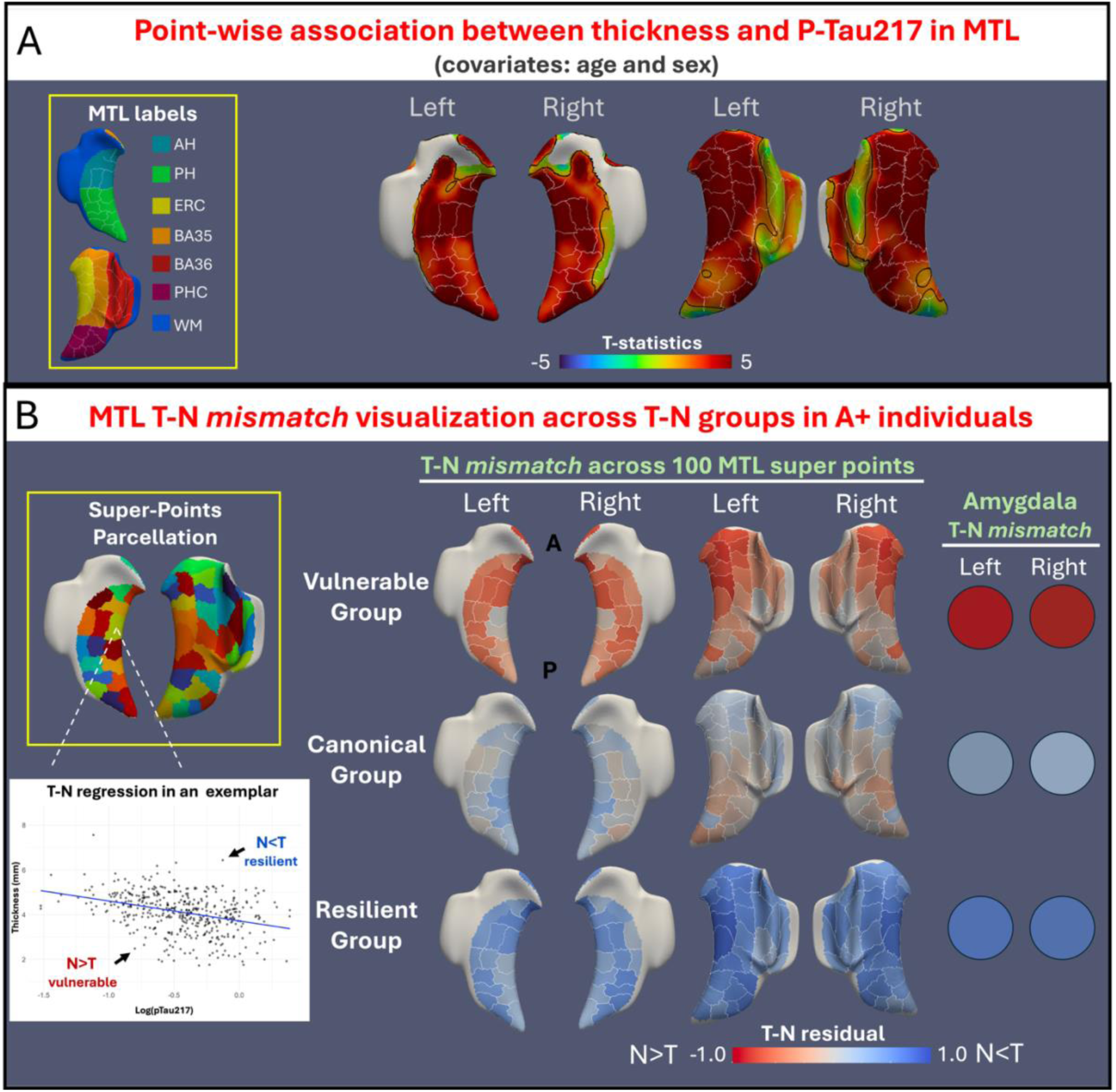
(**A**) Vertex-wise association between MTL thickness and p-tau_217_ for A+ individuals from ADNI after adjusting for age and gender. The white edges represent the outline of parcellated super points. The black edges indicate the area with significant association *P<0.05*. (**B**) Visualization of T-N residual patterns, calculated as standardized residuals (residuals divided by their standard deviation), across 100 super-points within the MTL and amygdala for A+ individuals.

We next performed T-N *mismatch* clustering based on deviation from the relationship of ptau_217_ with cortical thickness from 100 MTL super-points and amygdala volume, identifying three T-N subgroups. The “canonical” group (32.4%) displayed minimal T-N residuals across MTL and amygdala, reflecting a relatively close alignment between tau burden and neurodegeneration (T∼N). The “vulnerable” group (36.9%) exhibited negative T-N residuals, with greater thinning than expected based on ptau_217_ levels, particularly in the amygdala and anterior MTL regions (N > T). In contrast, the “resilient” group (30.6%) displayed positive T-N residuals, suggesting relative preservation of structure despite elevated tau, most prominently in the amygdala and diffuse extrahippocampal areas (T > N). These groups were relatively balanced in size and exhibited distinct patterns of T-N mismatch (Figure 1B). Significant difference in ptau_217_ was observed with the vulnerable group showing higher ptau_217_ level than the canonical group (p=0.039, Table 1). All subsequent analyses were adjusted for ptau_217_ to account this difference.

**Table 1.** Comparisons of demographic and other characteristics between T-N groups for A+ individuals (n=447) from ADNI. Overall group effects (p-values indicated in the bottom row) were tested using the Kruskal-Wallis test for categorical variables and linear regression for continuous variables. Covariates for comparisons are listed in the table. Only significant pairwise comparisons between vulnerable and resilient groups with the canonical group are marked in the table. All comparisons were two-sided. The p-values were Bonferroni-corrected for multiple comparisons (\**p<0.05, **p<0.01, ***p<0.001*). Abbreviations: **AH**=Anterior Hippocampus, **PH**=Posterior Hippocampus, **ERC**=Entorhinal Cortex, **PHC**= Parahippocampal Cortex, **Amyg**=Amygdala, **MEM**=Memory, **LAN**=Language, **EXF**=Executive Function, Dem = dementia.

| ADNI dataset |  |  |  |  |  |  |  |  |  |  |  |  |
| --- | --- | --- | --- | --- | --- | --- | --- | --- | --- | --- | --- | --- |
| Description | Basic Characteristics<br>Covariates for $\text{ptau}_{217}$: age, and sex | | | | Imaging Features<br>Covariates: $\text{ptau}_{217}$, age, sex and ICV (ICV not included for ERC/PHC) | | | | Cognition<br>Covariates: $\text{ptau}_{217}$, age, sex and years of education | | | |
| | Age (yrs) | Sex (F/M) | $\text{ptau}_{217}$ | Diagnosis (CU/ MCI/ Dem) | AH Vol (mm<sup>3</sup>) | PH Vol (mm<sup>3</sup>) | ERC/ PHC Ratio | Amyg Vol (mm<sup>3</sup>) | CDRSB | MEM | LAN | EXF |
| Group1<br>Vulnerable<br>(N>T)<br>(n=165) | 77.6<br>(7.6) | 85/80 | 0.509<br>(0.39)<br>* | 25/<br>56/<br>84 | 1415<br>(274)<br>*** | 1359<br>(284)<br>*** | 0.923<br>(0.089)<br>*** | 964<br>(167)<br>*** | 4.09<br>(3.6)<br>*** | -0.46<br>(0.9)<br>*** | -0.050<br>(0.7)<br>*** | -0.15<br>(0.8)<br>*** |
| Group2<br>Canonical<br>(N~T)<br>(n=145) | 76.2<br>(7.3) | 73/72 | 0.409<br>(0.36) | 72/<br>47/<br>26 | 1689<br>(249) | 1554<br>(168) | 1.04<br>(0.081) | 1183<br>(136) | 1.61<br>(2.8) | 0.37<br>(0.9) | 0.47<br>(0.8) | 0.35<br>(0.8) |
| Group3<br>Resilient<br>(N<T)<br>(n=137) | 74.3<br>(7.9) | 62/75 | 0.504<br>(0.39) | 66/<br>56/<br>15 | 1814<br>(249)<br>*** | 1359<br>(284)<br>*** | 1.01<br>(0.079) | 1236<br>(137)<br>** | 1.05<br>(1.5)<br>** | 0.51<br>(0.7)<br>** | 0.55<br>(0.6)<br>* | 0.33<br>(0.7) |
| Group Diff. | $P<0.01$ | $P=0.53$ | $P=0.039$ | $P<0.001$ | $P<0.001$ | $P<0.001$ | $P<0.001$ | $P<0.001$ | $P<0.001$ | $P<0.001$ | $P<0.001$ | $P<0.001$ |

The demographics of these T-N groups is shown in Table 1. The groups differed in age (*p<0.05*), with the vulnerable group tending to be older than the canonical and resilient group. There were no differences in sex distribution across groups (*p=0.53*).

### Cross-sectional and longitudinal atrophy differences between T-N groups

We next compared structural differences among ADNI T-N groups. While we did observe some increased ptau_217_ level in the vulnerable and resilient group relative to canonical (*p=0.039*), we observed significant difference in regional structure (Table 1) after controlling ptau_217_, age, sex and ICV. The vulnerable group had significantly reduced amygdala volume compared to the canonical group (*p<0.001*) while the resilient group had larger volume (*p<0.01*). The vulnerable group displayed significantly greater atrophy in both anterior and posterior hippocampal regions (*p<0.001*) whereas the resilient group demonstrated greater volume (*p<0.001*). Additionally, the T-N groups differed in the entorhinal cortex to parahippocampal cortex ratio (ERC/PHC) which is an imaging feature suggestive of comorbid LATE in AD, with the vulnerable group showing a significantly lower ratio than the others (*p<0.001*).

To further characterize structural differences, we performed vertex-wise cortical thickness comparison across the MTL (Figure 2A). In line with T-N *mismatch* patterns and regional volume results, the vulnerable group exhibited significantly greater atrophy than the canonical group, prominently in the anterior extrahippocampal and hippocampal regions within the MTL. In contrast, the resilient group displayed greater thickness in widespread MTL regions. Extending to the whole-brain comparisons (Figure 2B), these patterns were further supported, with the vulnerable group showing anterior temporal-limbic and orbito-frontal thinning relative to the canonical group, while the resilient group exhibited preservation of cortical thickness in temporal lobe and posterior cortices.

**Figure 2.**
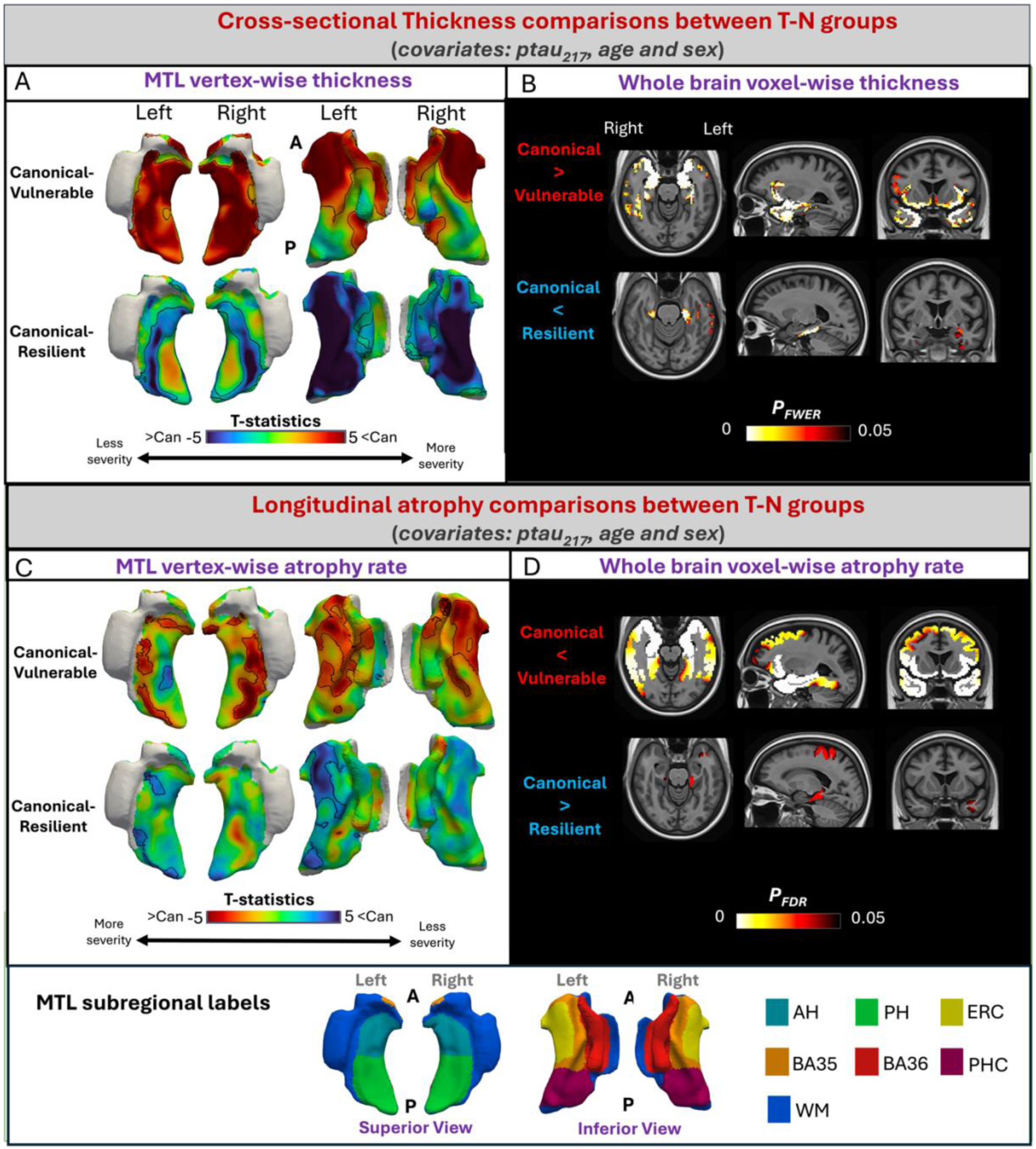
(**A**) T statistics for vertex-wise thickness comparisons: canonical versus vulnerable group (canonical – vulnerable) in the top row and canonical versus resilient group (canonical – resilient) in the bottom row. Comparisons were adjusted for p-tau_217_, age, and sex. Positive T-values indicates less thickness for vulnerable/resilient group compared with the canonical group. The black edges highlight the area with significant thickness comparisons *P_FWER_<0.05*. **(B)** Voxel-wise thickness comparisons in whole brain between the vulnerable and canonical groups (top) and between resilient and canonical groups (bottom). Comparisons were adjusted by ptau_217_, age and sex. The areas with black outline represent significant differences for the vulnerable and resilient groups with the canonical group (*P_FWER_ < 0.05*). **(C)** T-statistics for vertex-wise longitudinal atrophy comparisons using linear mixed-effects models: canonical vs. vulnerable (top) and canonical vs. resilient (bottom), adjusted for ptau_217_, age, and sex. Negative T-values indicate greater atrophy in the comparison group relative to the canonical group. The black edges represent significant differences with the canonical group for each T-N *mismatch* group (*P_FDR_ < 0.05*). **(D)** Voxel-wise longitudinal cortical thickness changes across the whole brain using linear mixed-effects models: canonical vs. vulnerable (top) and canonical vs. resilient (bottom), adjusted for ptau_217_, age, and sex. Black edge outlines indicate areas with significant differences in atrophy rates compared to the canonical group (*P_FDR_ < 0.05*).

Longitudinal results were consistent with cross-sectional patterns (Figure 2C-D). Compared to the canonical group, the vulnerable group displayed greater atrophy over time, particularly in the anterior extrahippocampal and anterior temporal regions. In contrast, the resilient group exhibited slower cortical thinning relative to the canonical group.

In addition to the comparisons between T-N groups, we also compared the structural differences of the ADNI T-N groups with A-CU (Figure S2). The vulnerable group exhibited the most extensive atrophy, both within the MTL and across the whole brain. The canonical group showed a more uniform pattern of atrophy within the MTL, along with AD-like posterior cortical thinning. The resilient group demonstrated only focal significant changes both cross-sectionally and longitudinally, with a mild AD-like atrophy pattern.

### The vulnerable group exhibited early atrophy prior to predicted onset time of Tau positivity

We next examined longitudinal structural changes up to 15 years preceding estimated Tau onset across T-N groups in ADNI (Figure 3A-B). We observed that the vulnerable group displayed significantly greater atrophy in anterior extrahippocampal regions of the MTL and broader limbic areas across the brain, compared to the canonical group. The resilient group exhibited no significant differences to the canonical group (Figure 3A–B).

**Figure 3.**
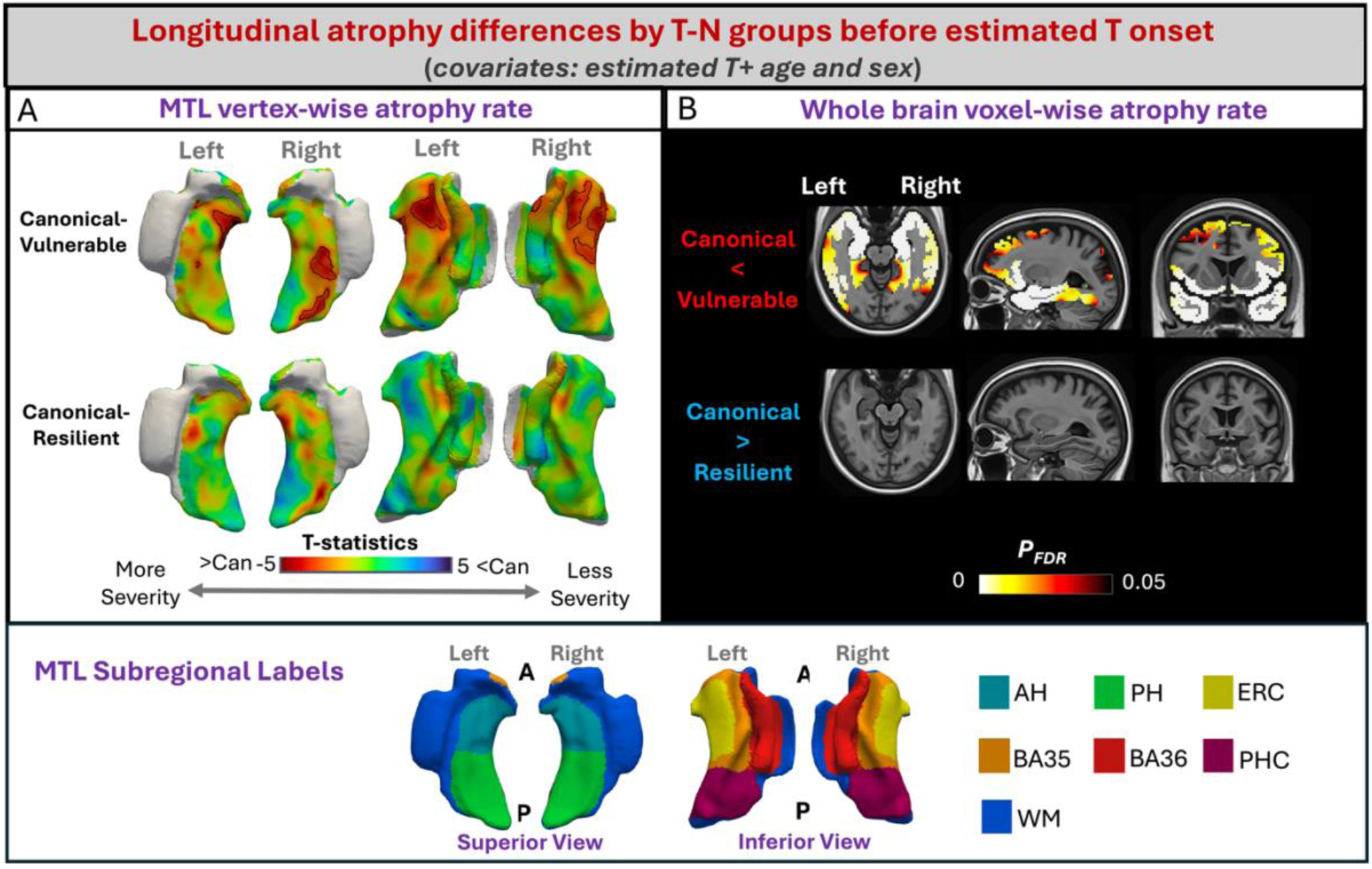
**(A)** Vertex-wise T-maps comparing pre-Tau onset atrophy between canonical and mismatch groups (vulnerable, top; resilient, bottom), adjusted for estimated Tau onset age and sex. Negative T-values indicate greater atrophy relative to the canonical group. The black edges highlight the area with significant thickness comparisons *P_FDR_<0.05.* **(B)** Voxel-wise comparisons of whole-brain cortical thinning before Tau positivity between groups (*P_FDR_ < 0.05*).

### T-N groups displayed distinct cognitive profiles

At baseline, the vulnerable group displayed significantly higher CDRSB score (*p<0.001*) compared with the canonical group, whereas the resilient group had a lower (*p<0.01*) mean CDRSB measure (Table 1). The groups also differed in domain-specific cognitive functions. The vulnerable group displayed significantly worse performance in memory, language and executive function relative to the canonical group (*p<0.001*). In contrast, the resilient group demonstrated better performance in memory (*p<0.01*) and language (*p<0.05*).

We further evaluated longitudinal cognitive trajectories relative to estimated T+ onset age across the T-N groups. Change in CDRSB, memory, language, and executive function over time were modeled after adjusting for age at T+ onset, sex, and years of education (Figure 4A). The vulnerable group exhibited significantly faster increase in CDRSB scores compared to the canonical group (*p < 0.001*). At the domain level, the vulnerable group demonstrated greater decline in memory (*p < 0.001*), language (*p < 0.01*), and executive function (*p < 0.001*) relative to the canonical group although with greatest differences in memory in absolute terms. Notably, the vulnerable group showed signs of cognitive impairment, especially in memory, prior to estimated T+ (Figure 4). We did not observe significant difference between the resilient and the canonical group in cognitive trajectories. We also modeled longitudinal cognitive trajectories anchored to baseline rather than T+ time and observed similar patterns (Figure S3).

**Figure 4.**
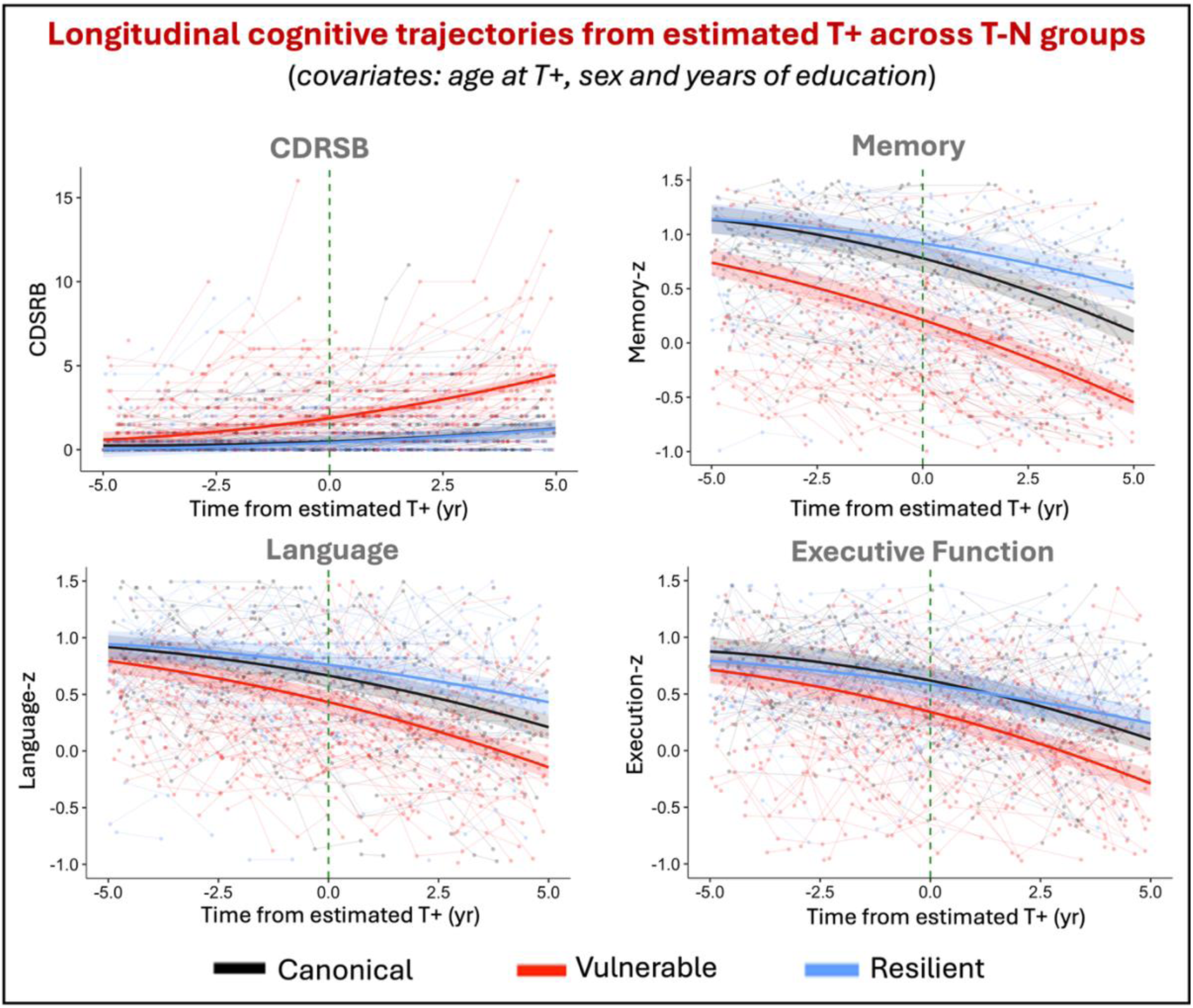
Longitudinal cognitive trajectories aligned to estimated T+ across CDRSB, memory, language and executive function z-scores between T-N groups. Shaded ribbons indicate 95% confidence intervals for model-predicted trajectories, while semi-transparent spaghetti lines represent individual individual-level cognitive trajectories over time.

### T-N groups were replicated in an independent cohort

To evaluate the robustness of our approach, we replicated the analysis in an independent cohort of A+ individuals, including cognitively unimpaired, MCI and dementia-level impairment (Table S1) from the Penn ADRC. Once again, we identified three distinct T-N groups (Figure 6A): vulnerable (N>T, 12%), canonical (N∼T, 53.7%), and resilient (N<T, 34.3%). The vulnerable group, similar to in the ADNI cohort, displayed disproportionate neurodegeneration relative to tau burden in the amygdala, hippocampus, and anterior extrahippocampal areas (Figure 5A). In contrast, the resilient group displayed neurodegeneration less than expected for T in a broad pattern across the MTL and the canonical group had low T-N residuals across MTL super-points and the amygdala.

**Figure 5.**
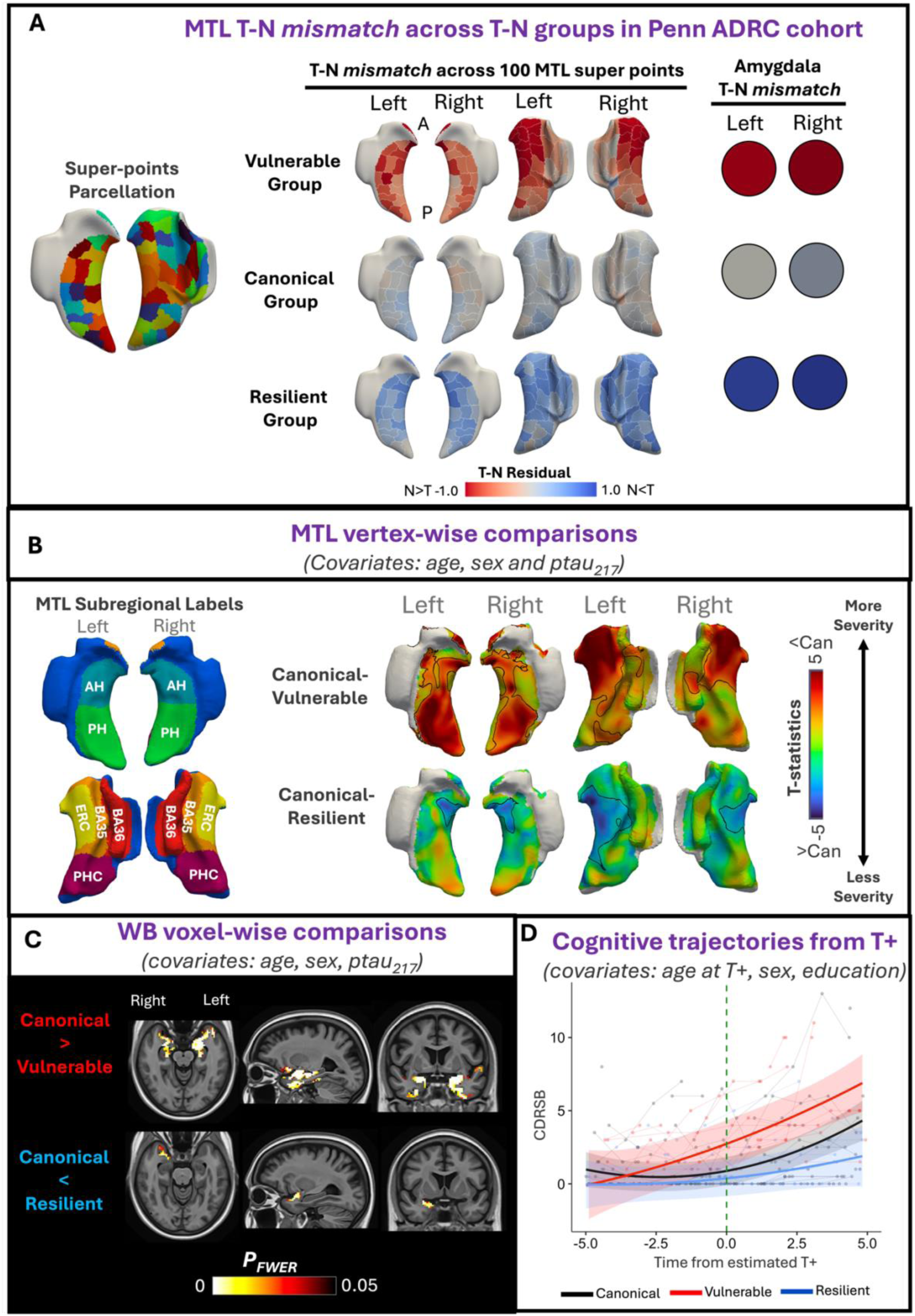
Replication of MTL T-N *mismatch* from Penn ADRC dataset. **(A)** Visualization of T-N residual patterns across 50 bilateral super-points within the MTL and amygdala for 108 A+ individuals from the Penn ADRC. The white edges represent the outline of parcellated super points. **(B)** T statistics for vertex-wise thickness comparisons among T-N groups from Penn ADRC cohort: canonical versus vulnerable group (canonical – vulnerable) in the top row and canonical versus resilient group (canonical – resilient) in the bottom row. Comparisons were adjusted for p-tau_217_, age, and sex. Negative T-values indicates less thickness for vulnerable/resilient group compared with the canonical group. The black edges highlight the area with significant thickness comparisons *P_FWER_<0.05*. **(C)** Voxel-wise whole brain thickness comparisons between the vulnerable group and the canonical group (top) and between the resilient group and the canonical group (bottom). Comparisons were adjusted by ptau_217_, age and sex. The colored areas represent significant differences with the canonical group for each T-N mismatch group (*P_FWER_ < 0.05*). WB= whole brain **(D)** Longitudinal cognitive trajectories relative to estimated T+ for CDRSB between T-N groups. Shaded ribbons indicate 95% confidence intervals for model-predicted trajectories, while semi-transparent spaghetti lines represent individual participant-level cognitive trajectories over time.

**Figure 6.**
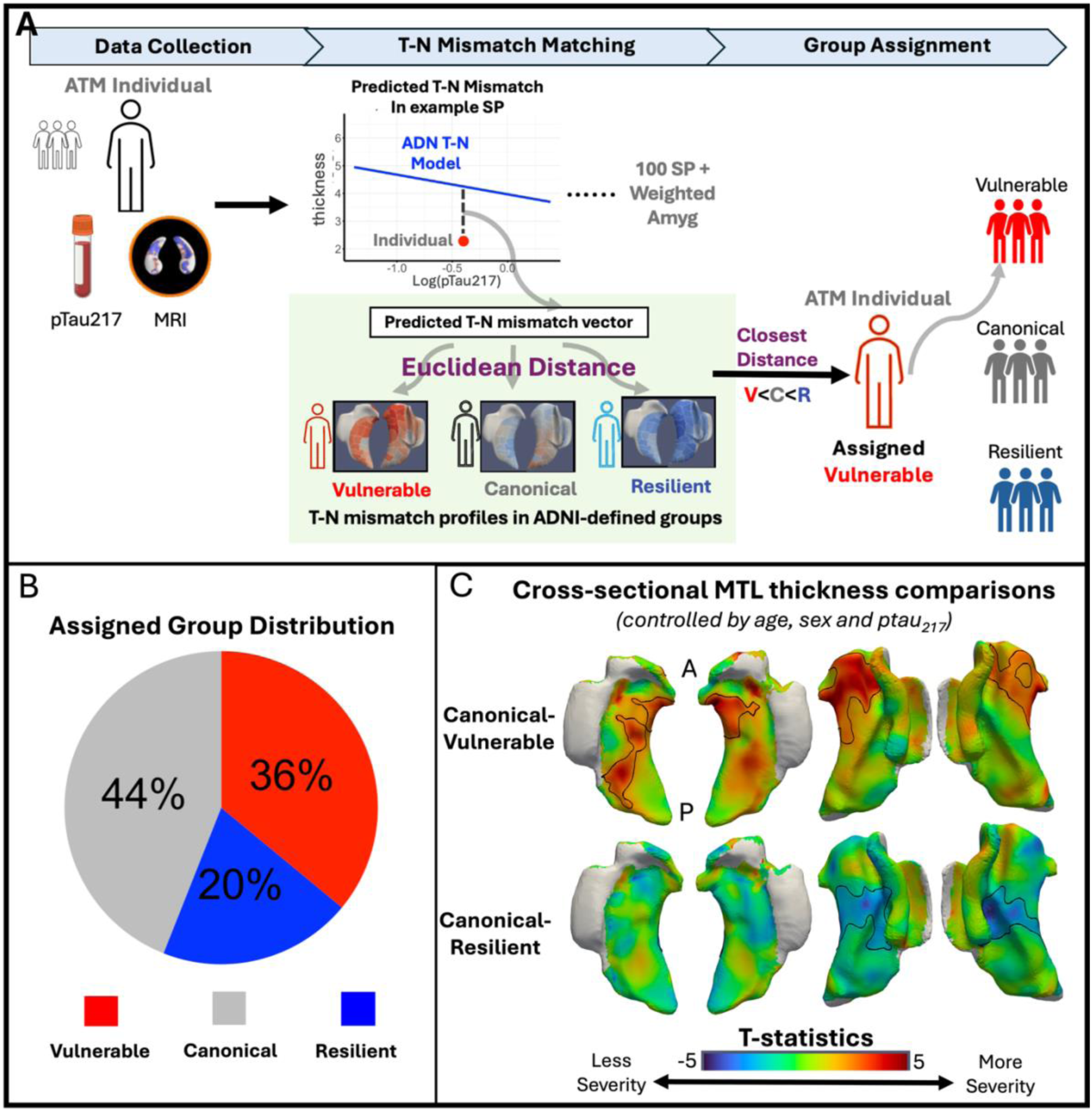
Application of MTL T-N mismatch model to 50 patients treated with anti-amyloid therapeutics. **(A)** Schematic illustrating group assignment for a representative individual based on T–N mismatch. Abbreviations: SP=Super-Points, V=Vulnerable, C=Canonical, R=Resilient. **(B)** The pie chart showing the distribution of assigned T-N groups among the 50 individuals. **(C)** Vertex-wise thickness comparisons between T-N groups in the ATM cohort: canonical vs. vulnerable (top) and canonical vs. resilient (bottom), adjusted for p-tau_217_, age, and sex. Negative T-values indicate lower thickness in vulnerable/resilient groups. Black outlines denote regions with significant differences (*P_FWER_ < 0.05*).

Consistent with findings from the ADNI cohort, the vulnerable group in the Penn ADRC cohort exhibited a lower ERC/PHC ratio (*p<0.001*), reduced amygdala volume (*p<0.001*), and greater hippocampal atrophy (Table S2). Within the MTL, this group displayed significantly reduced cortical thickness in both the hippocampus and specifically anterior extrahippocampal regions compared to the canonical group (Figure 5B). Whole-brain analysis further revealed more extensive atrophy in anterior temporal-limbic regions (Figure 5C). The resilient group showed more thickness preservation in extrahippocampal region. Also similar to the ADNI cohort, the vulnerable group tended to exhibit higher baseline CDRSB scores and faster longitudinal cognitive decline (Figure 5D), although this did not reach statistical significance.

### Applicability of T–N Mismatch to Clinical Practice

Finally, we evaluated the applicability of this approach in a real-world clinical cohort (ATM) of 50 AD patients who underwent anti-amyloid therapy. A schematic illustrating the workflow for an exemplar patient is shown in Figure 6A. Using the T–N models derived from ADNI, we predicted individual T–N mismatch profiles across MTL super-points and amygdala in the ATM cohort. Each patient was then assigned to the most similar T–N phenotype by computing the Euclidean distance between their residual pattern and the centroid of each ADNI-derived cluster and selecting the phenotype corresponding to the closest distance. After assignment (Figure 6B), 44% of patients were classified as canonical (n=22), 36% as vulnerable (n=18), and 20% as resilient (n=10). This distribution was similar to the T-N group proportions observed in the ADNI cohort. We then examined group-level characteristics following this assignment (Table S3). Despite the vulnerable group having lower ptau_217_ levels than the canonical group (*P<0.01*), it demonstrated lower volumes in the amygdala (*P<0.001*) and hippocampus (*P<0.001*)—especially in the anterior hippocampus—compared to the canonical group, after adjusting for ptau_217_, age, sex, and ICV. In contrast, the resilient group showed higher hippocampal volume (*P<0.05*). Differences were also observed in the ERC/PHC ratio (*P<0.01*), with the vulnerable group exhibiting a lower ratio (*P<0.05*). Furthermore, vertex-wise comparisons revealed greater atrophy in the anterior extrahippocampal regions in the vulnerable group relative to the canonical group (Figure 6C), whereas the resilient group showed less atrophy. These findings closely align with those observed in the ADNI-defined T– N groups and the Penn ADRC replication cohort.

## Discussion

Alzheimer’s disease is heterogeneous, with neurodegeneration often influenced by co-pathologies and resilience factors. Given the current limitations of *in vivo* biomarkers for detecting co-pathologies, indirect approaches to infer their presence and severity are valuable for prognosis and potentially for determining appropriate patients for targeted therapies. We demonstrated that *mismatch* between tau and neurodegeneration across the whole brain identified individuals who likely harbor co-pathologies (e.g., TDP-43, vascular burden) that may contribute to greater vulnerability and faster rate of cognitive decline. Additionally, this approach also classified some individuals as resilient with a more indolent rate of decline, as shown previously^29–31^.

The current analysis focused specifically on the MTL, a region associated with not only the early neurofibrillary tangle (NFT) pathology of AD, but also of one of its most common co-pathologies, LATE-NC, which is present in up to 50% of AD patients^56^. Previous work has demonstrated that when AD co-occurs with LATE-NC, patients have a faster rate of cognitive decline and hippocampal atrophy^10,57–59^. Thus, identification of such patients has particular clinical significance in practice and clinical trials. LATE-NC does not uniformly affect the MTL and has a spatial pattern that does not completely overlap with typical AD. Specifically, LATE-NC displays an anterior-to-posterior gradient of atrophy, including early involvement of the amygdala (Stage 1), the anterior hippocampus and the entorhinal cortex (Stage 2) ^19,21,24^. In contrast, typical AD tends to involve the posterior MTL to a greater extent than LATE, particularly the parahippocampal cortex^19^, although both AD and LATE exhibit a degree of predilection for the anterior MTL^10,36^. To capture these spatial differences, we parcellated the MTL into 100 super-points and incorporated the amygdala for volume-based weighted T-N *mismatch* clustering.

To potentially increase the clinical accessibility of the approach described, we used plasma ptau_217_, as a global measure of tau pathology, as opposed to Tau PET. The latter provides important spatial information, but is currently not generally available in clinical practice, limited by variable insurance coverage, cost, and access to PET scanners. Plasma measures offer a much less expensive alternative. Previous studies^60,61^ demonstrated that while plasma ptau_217_ is highly sensitive to early amyloid-β status, it also has strong correlation with both temporal and global measures of tau burden measured by Tau PET.

Using this approach, we identified three data-driven T-N groups that were labelled as vulnerable (N>T), canonical (N∼T), or resilient (N<T) among patients on the AD continuum. The vulnerable group displayed a number of features suggestive of the presence of LATE-NC, including more severe atrophy in anterior MTL regions, as evidenced by a lower ERC/PHC ratio, a structural signature previously found to be associated with AD with concomitant TDP-43 in postmortem studies^19^. Additionally, this group displayed a pattern of atrophy outside the MTL in anterior temporal-limbic areas and fronto-insular and orbitofrontal cortices that reflect regions of TDP-43 progression in LATE-NC^59,62^. Longitudinal patterns of atrophy largely overlapped with the cross-sectional findings.

In addition to the imaging features suggesting concomitant LATE-NC in the vulnerable group, its cognitive profile was also consistent with studies of mixed AD and LATE-NC. In particular, memory impairment is the hallmark clinical feature of LATE-NC, and semantic memory/language may also be involved as well^10,56,63^. Consistent with this pattern and a faster rate of decline than typical AD, the vulnerable group displayed most prominently a faster rate of decline in memory followed by language and executive measures.

Interestingly, the vulnerable group showed LATE-like atrophy patterns even before reaching estimated T+, as well as evidence of progressive memory impairment prior to T+. In light of strong relationship between development of tau pathology with cognitive decline^64–66^, this finding suggests an independent, non-AD process also contributing to decline in this group providing further support for LATE-NC as a contributor in these cases. It also suggests that some cases that are A+ may have LATE-NC as an earlier driver of symptoms prior to the evolution of AD pathology^67^.

In contrast, the resilient group, defined by less neurodegeneration than expected given their tau burden, showed greater preservation of thickness and slower rates of atrophy in the MTL and whole brain. In our prior work^29–31^ examining whole-brain T-N *mismatch*, younger age was found to be a potential protective factor associated with the resilient phenotype. However, in the present study, where resilience was defined based on MTL-specific T-N mismatch, only a modest age difference was observed, suggesting that distinct or additional protective mechanisms may be involved. One possibility is that these individuals may be particularly free from co-pathologies while the canonical group may harbor an intermediate burden. Future research is needed to better understand the underlying factors contributing to resilience in this group.

We reproduced the T-N groups and observed consistent structural and cognitive patterns across groups in an independent research dataset. The vulnerable group again demonstrated features suggestive of LATE-NC involvement, including reduced ERC/PHC ratio, lower amygdala volume, and anterior MTL atrophy, along with accelerated cognitive decline. The resilient group remained cognitively preserved despite tau burden. These replicated findings support the potential utility of T-N *mismatch* modeling as a reliable and generalizable in vivo approach for identifying susceptibilities and resilience within the AD continuum.

Importantly, we extended this approach to a real-world clinical cohort of AD patients undergoing anti-amyloid therapy, representing the first application of this T–N mismatch framework beyond research cohorts. The observation of similar T–N group patterns in this clinical dataset supports the generalizability and translational potential of this approach. Among the 50 treated AD patients, only 44% were classified as canonical, while the remaining 56% exhibited divergent T–N mismatch patterns, including 36% identified as vulnerable and 20% as resilient. Consistent with the findings from ADNI, vulnerable individuals exhibited a lower ERC/PHC ratio compared to the canonical and resilient groups, with the value similar to what observed in the ADNI vulnerable group. These findings highlight the potential of the approach for disentangling the heterogeneity in treatment populations, and it will be important to determine how these groups respond to such treatments in this context. For example, if the canonical or resilient group harbor less co-pathology, they may show the most robust effects of an amyloid specific therapy.

Our work has limitations. The choice of parameters, such as the number of super-points and the number of T-N groups, was made empirically. Second, we used clinically accessible plasma ptau_217_ as a measurement for T. While previous studies have shown that ptau_217_ correlates strongly with tau PET, it also reflects amyloid burden and does not provide spatially specific information. Third, while the vulnerable group exhibited imaging and clinical features suggestive of LATE-NC involvement, we lack pathological confirmation. Future postmortem studies are needed to further validate this association. Fourth, the primary T–N mismatch modeling was based on the ADNI cohort, which may not reflect the full spectrum of clinical populations. ADNI participants are generally less diverse in terms of race, ethnicity, and disease severity compared to real-world patients. In the included datasets, 89% of participants in ADNI and 87% in the Penn ADRC identified as white. As such, broader validation across more heterogeneous cohorts is necessary to ensure the robustness and clinical applicability of this approach for the future work.

## Conclusion

In conclusion, T-N *mismatch* within the MTL using MRI and plasma biomarkers revealed groups with varying vulnerability/resilience, with the vulnerable group displaying patterns of atrophy and cognition suggestive of LATE-NC. This approach provides important prognostic information and is a potentially cost-effective method for stratifying individuals for therapeutic interventions. Such stratification will likely be important in current and future interventions given the heterogeneity of AD, which requires a precision medicine-based approach. To support broader use, we released a public software package (https://github.com/xueying-lyu/mismatch) implementing user-customized mismatch clustering and individual-level T-N phenotypic assignment as developed in this study.

## Data Availability

All data produced in the present study are available upon reasonable request to the authors

## CONFLICTS

D.A.W has served as a paid consultant to Eli Lilly, GE Healthcare, and Qynapse. He serves on a DSMB for Functional Neuromodulation. He is a site investigator for a clinical trial sponsored by Biogen. I.M.N serves as speakers bureau for Biogen and advisory board for Eisai. L.X. received personal consulting fees from Galileo CDS, Inc. He has become an employee of Siemens Healthineers since May 2022 but the current study was conducted during his employment at the University of Pennsylvania.

## ACKNOWLEDGEMENT/FUNDING

The study was supported by the funding R01 AG056014 and RF1 AG069474. LW is supported by National Institute on Aging grant R01AG070592, MultiPark - A Strategic Research Area at Lund University, the Bente Rexed Gerstedt Foundation and the Swedish Research Council (2022-00900). Data collection and sharing for this project was funded by the Alzheimer’s Disease Neuroimaging Initiative (ADNI) (National Institutes of Health Grant U01 AG024904) and DOD ADNI (Department of Defense award number W81XWH-12-2-0012).

Data collection and sharing for this project was funded by the Alzheimer’s Disease Neuroimaging Initiative (ADNI) (National Institutes of Health Grant U01 AG024904) and DOD ADNI (Department of Defense award number W81XWH-12-2-0012). ADNI is funded by the National Institute on Aging, the National Institute of Biomedical Imaging and Bioengineering, and through generous contributions from the following: AbbVie, Alzheimer’s Association; Alzheimer’s Drug Discovery Foundation; Araclon Biotech; BioClinica, Inc.; Biogen; Bristol-Myers Squibb Company; CereSpir, Inc.; Cogstate; Eisai Inc.; Elan Pharmaceuticals, Inc.; Eli Lilly and Company; EuroImmun; F. Hoffmann-La Roche Ltd and its affiliated company Genentech, Inc.; Fujirebio; GE Healthcare; IXICO Ltd.; Janssen Alzheimer Immunotherapy Research & Development, LLC.; Johnson & Johnson Pharmaceutical Research & Development LLC.; Lumosity; Lundbeck; Merck & Co., Inc.; Meso Scale Diagnostics, LLC.; NeuroRx Research; Neurotrack Technologies; Novartis Pharmaceuticals Corporation; Pfizer Inc.; Piramal Imaging; Servier; Takeda Pharmaceutical Company; and Transition Therapeutics. The Canadian Institutes of Health Research is providing funds to support ADNI clinical sites in Canada. Private sector contributions are facilitated by the Foundation for the National Institutes of Health (www.fnih.org). The grantee organization is the Northern California Institute for Research and Education, and the study is coordinated by the Alzheimer’s Therapeutic Research Institute at the University of Southern California. ADNI data are disseminated by the Laboratory for Neuro Imaging at the University of Southern California.

## Notes

### Competing Interest Statement

The authors have declared no competing interest.

### Author Declarations

Ethics committee/IRB of University of Pennsylvania name gave ethical approval for this work

