## Supplemental Materials for "Medial temporal lobe Tau-Neurodegeneration *mismatch* from MRI and plasma biomarkers identifies vulnerable and resilient phenotypes with AD"

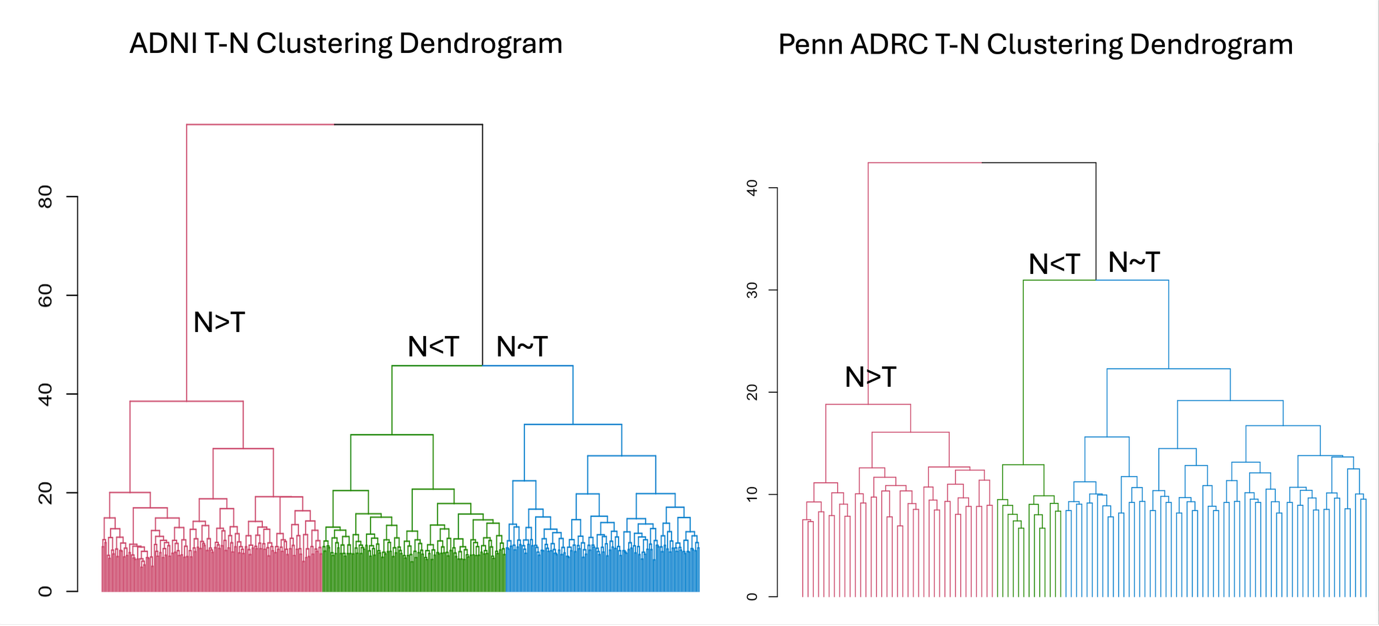

**Figure S1.** Hierarchical clustering dendrogram for ADNI T-N mismatch clustering (A) and Penn ADRC replication T-N mismatch clustering (B).

**Table S1.** Clinical characteristics of A+ subjects from ADNI, Penn ADRC and ATM

| Cohort | Age | Sex  (Female/  Male) | ptau_217_ | Diagnosis  (CU/  MCI/  Dementia/  unknown) | CDRSB |
| --- | --- | --- | --- | --- | --- |
| ADNI  (447) | 76.1  (7.7) | 220/227 | 0.475  (0.38) | 163/159/125/0 | 2.35  (7.7) |
| Penn ADRC  (n=108) | 74.6  (6.6) | 61/47 | 0.519  (0.41) | 39/36/32/1 | 1.84  (2.3) |
| ATM  (N=50) | 72.5  (8.0) | 27/23 | 0.66  (0.44) | 1/29/20/0 |  |

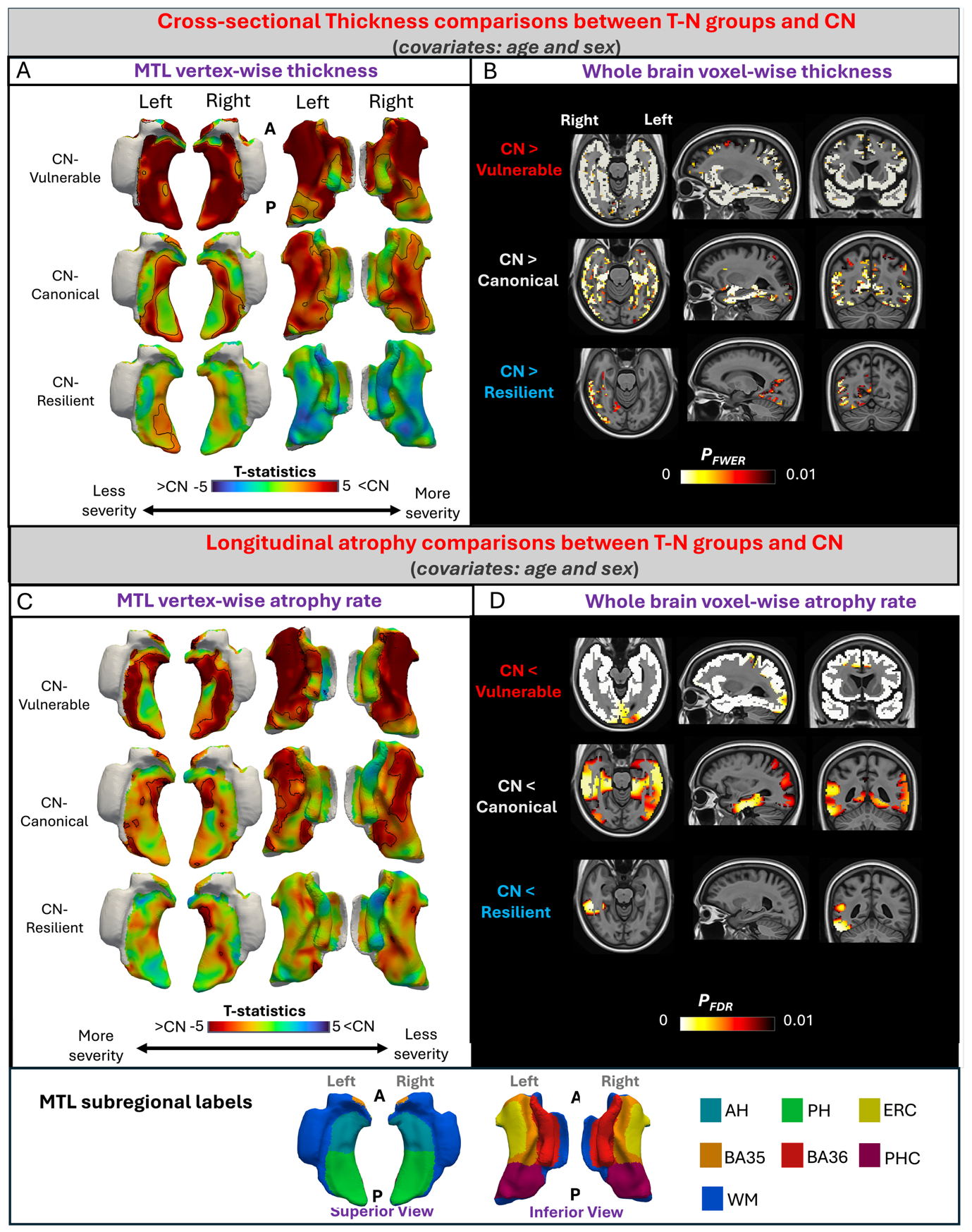

**Figure S2. (A)** T-statistics for point-wise cortical thickness comparisons between controls (CU) and each T–N group (vulnerable, canonical, resilient), adjusted for age and sex. Positive values indicate reduced thickness in T–N groups. Black outlines denote significant regions (*P_FWER_ < 0.01*). **(B)** Voxel-wise cortical thickness comparisons across the whole brain between controls and each T–N group (*P_FWER_ < 0.01*), adjusted for age and sex. **(C)** T-statistics for longitudinal point-wise atrophy comparisons using linear mixed-effects models, adjusted for age and sex. Negative values indicate greater atrophy in T–N groups (*P_FDR_ < 0.01*). **(D)** Voxel-wise longitudinal cortical thinning differences between controls and each T–N group (*P_FDR_ < 0.01*), adjusted for age and sex.

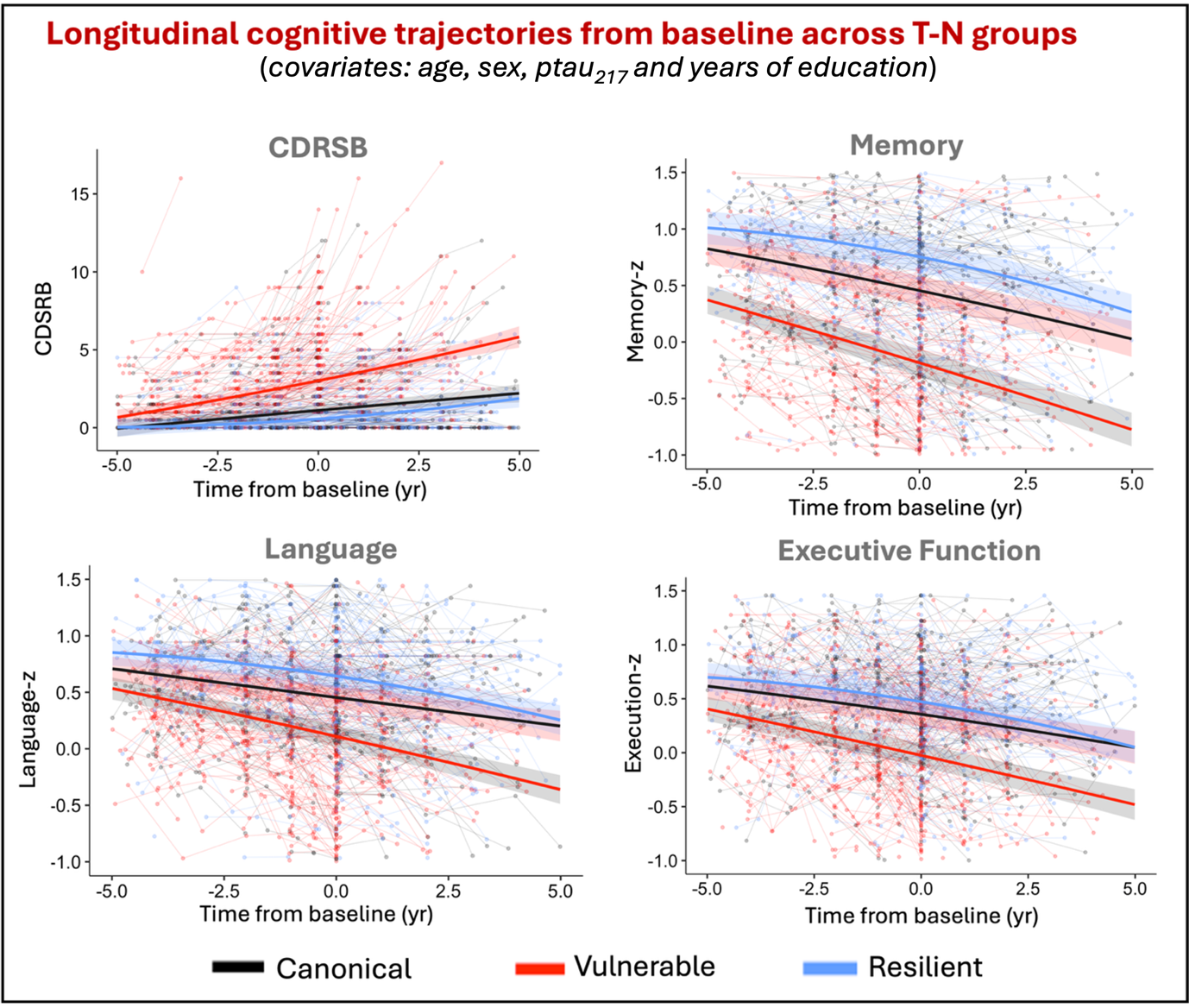
**Figure S3.** Longitudinal cognitive trajectories from baseline across CDRSB, memory, language and executive function z-scores between T-N groups. Shaded ribbons indicate 95% confidence intervals for model-predicted trajectories, while semi-transparent spaghetti lines represent individual individual-level cognitive trajectories over time.

**Table S2**. Comparisons of clinical features between T-N groups from Penn ADRC cohort (n=108). Overall group effects (p-values indicated in the bottom row) were tested using the Kruskal-Wallis test for categorical variables and linear regression for continuous variables. The comparison of ptau_217_ was controlled by age and sex. The covariates for CDRSB comparison included age, sex and ptau_217_. The covariates for imaging volumes or feature comparisons included age, ptau_217_, sex and ICV. Only significant pairwise comparisons between T-N groups and canonical group are marked in the table alongside the corresponding value. P-values were adjusted by Bonferroni multiple comparison correction (*P<0.05, **P<0.01, ***P<0.001). Abbreviations: AH=Anterior Hippocampus, PH=Posterior Hippocampus, ERC=Entorhinal Cortex, PHC= Parahippocampal Cortex

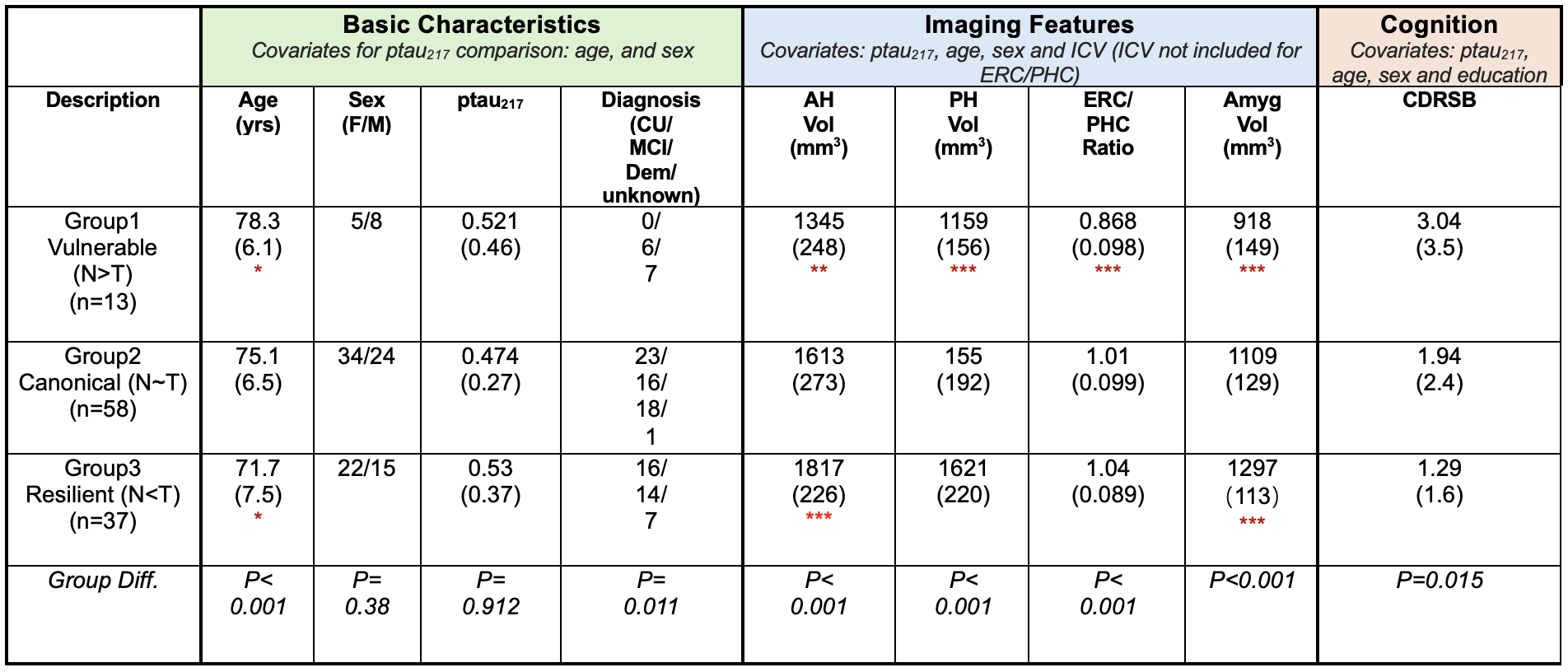

**Table S3**. Comparisons of clinical features between Assigned T-N groups from ATM cohort (n=50). Overall group effects (p-values indicated in the bottom row) were shown below. The comparison of ptau_217_ was controlled by age and sex. The covariates for CDRSB comparison included age, sex and pTau217. The covariates for imaging volumes or feature comparisons included age, ptau_217_, sex and ICV. Only significant pairwise comparisons between T-N groups and canonical group are marked in the table alongside the corresponding value. P-values were adjusted by Bonferroni multiple comparison correction (*P<0.05, **P<0.01, ***P<0.001). Abbreviations: AH=Anterior Hippocampus, PH=Posterior Hippocampus, ERC=Entorhinal Cortex, PHC= Parahippocampal Cortex

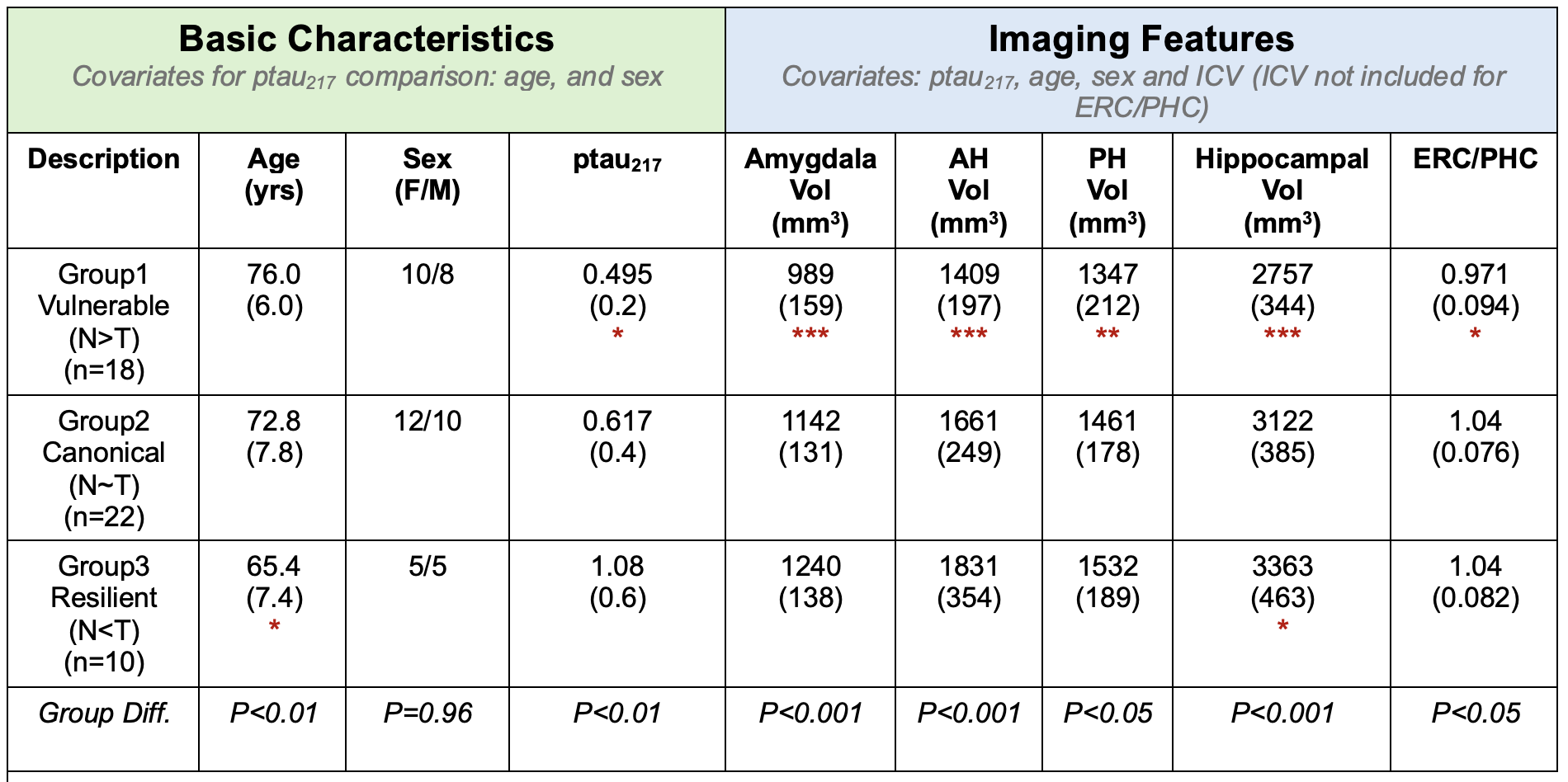
